# Profile of tetralogy of Fallot at the cardiology clinic of Saint-Damien hospital from January 2019 to December 2021

**DOI:** 10.1101/2023.12.14.23299857

**Authors:** Naïka Paulemie Désir, Adonaï Aly Isaac Julien, Richcard Alexandre, Joanne Fevry, Taina Brice, Alexandra Noisette

## Abstract

**Objective:** Our work aimed to study the epidemiological and clinical aspects of Tetralogy of Fallot (TF) in the cardiology unit of the Saint-Damien Nos Petits Frères et Sœurs hospital in Haiti.

**Methodology:** This is a retrospective study of patients aged 0 to 18 years with Tetralogy of Fallot confirmed by echocardiography. Data were entered using an elaborated form and analyzed with Epi Info software version 7.2.2.16 and Microsoft Excel 2013.

**Results:** From January 2019 to December 2021, we identified 102 patients aged 0 to 18 years with TF. A male predominance was noted with a sex ratio M/F of 1.55. The average age of discovery of our cohort was 3 years and 7 months [0-16 years; standard deviation: 9.9]. The clinical signs were dominated by a systolic murmur (91.18%) followed by cyanosis (39.22%) and dyspnea (33.33%). On echocardiography, the regular form (76.47%) associated with the severe stenosis (83.33%) was remarkable. 20.59% of patients were able to benefit from surgery and the mortality rate was 14.94%. Patients were presenting at an advanced stage where they mostly needed surgical treatment to survive.

**Conclusion:** This study shows that the prevalence of TF is also high in developing countries. The heart murmur at the pulmonary focus and the cyanosis were more present on physical examination. Patients were presenting at an advanced stage where they mostly needed surgical treatment to survive. Preventive measures are needed to reduce the mortality rate.

## 1. Introduction

Congenital malformations are anatomical or functional alterations of an organ and/or system that occur in utero. They are a frequent cause of morbidity and mortality in newborns. In general, it is considered that about 2-3% of newborns present at least one malformation during physical evaluation (1). Congenital heart disease is the most common birth defect (2,3). Among the cyanotic congenital heart diseases, Tetralogy of Fallot (TF) is the most frequent and accounts for 7-10% of congenital heart diseases (2). Today due to advances in technology, the prevalence of TF in developed countries is well known. There are hardly any studies relating to this heart disease in Haiti. Hence the interest of studying the epidemiological profile of Tetralogy of Fallot at the cardiology clinic of the Saint-Damien Hospital Nos Petits Frères et Sœurs from January 2019 to December 2021.

## 2. Materials and Methods

### Type of study

This is a descriptive and retrospective study of patients aged 0 to 18 with Tetralogy of Fallot seen at the cardiology clinic of Saint-Damien Hospital Nos Petits Frères et Soeurs from January 2019 to December 2021.

### Study framework

Saint-Damien Hospital, a daughter structure of “*Nos Petits Frères et Soeurs (NPFS)*”, is a Catholic philanthropic institution dedicated almost entirely to child care. It is located in the commune of Tabarre 41, in the WEST department in Haiti. Its geographic coordinates are −72.25457 longitude and 18.48086 latitude. The code is 11234. The cardiology clinic at Saint-Damien Hospital began in November 2018. It takes care of all children aged 0 to 18, consults about ten children every day and serves practically all the country. For the year 2021, 1147 consultations were carried out in this unit. The staff includes: A chief medical officer (cardio pediatrician) and a nurse.

### Study population

The study population consists of all patients aged 0 to 16, seen at the cardiology clinic of Saint-Damien Hospital Nos Petits Frères et Soeurs with a diagnosis of Tetralogy of Fallot from January 1, 2019 to December 31, 2021.

### Inclusion and exclusion

Inclusion criteria: Any child seen at the cardiology clinic during the study period (2019-2021) with the diagnosis of Tetralogy of Fallot in his chart confirmed by echocardiography.

Exclusion criteria: Incomplete information, illegible records.

### Data collection and analyzes

Data were entered using a pre-elaborated form and then distributed as a grouping of frequency, mean, median and standard deviation, with Epi Info software version 7.2.2.16 and Microsoft Excel 2013.

## 3. Results

From January 2019 to December 2021, 2,010 patients were seen at the cardiology clinic at Saint-Damien Hospital Nos Petits Frères et Soeurs. Among the registered cases, we found 102 patients with a diagnosis of TF meeting the inclusion criteria, i.e. 5.1% of cases. Breakdown by year the 102 patients were collected over a 3-year period from 2019 to 2021. The greatest number of patients was diagnosed in 2019 then there was a drop which remained stable between 2020 and 2021.

**Figure 1.**
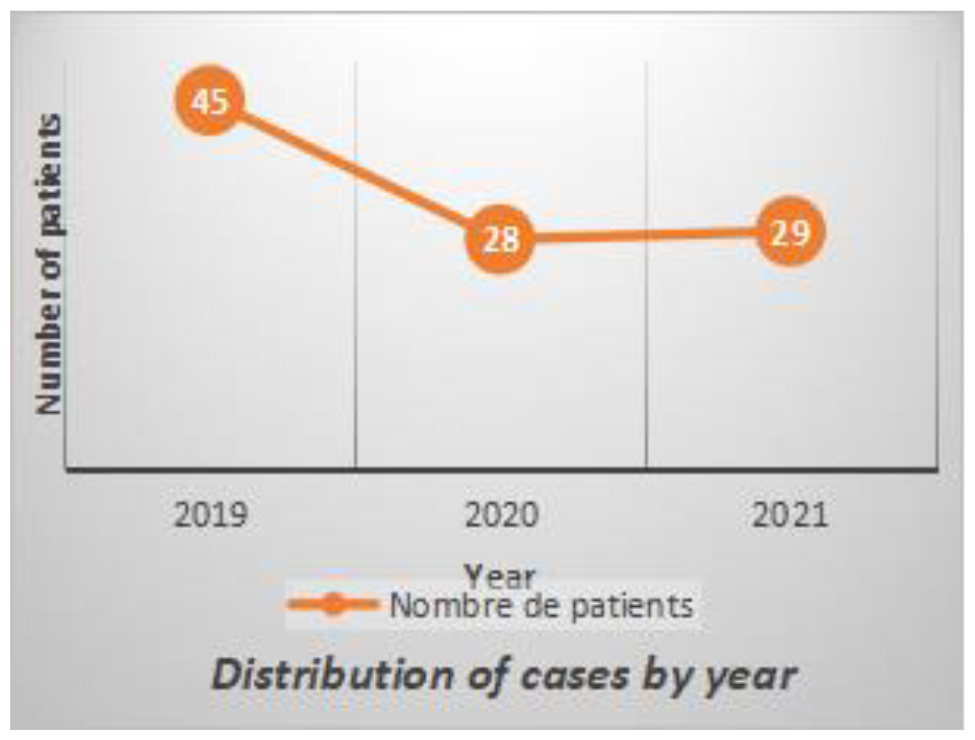
Distribution of cases by year.

### Breakdown of cases by age group

More than half of the cohort (69.61%) was diagnosed in the age groups of less than one year and 1 to 4 years. Among the less than one-year-old, there were 9 newborns.

### Measure of average age

The average age of the patients in the series was 3.7 years with a standard deviation of 9.9 and the extremes ranging from 0 to 16 years. The mode was less than 1 year and the median age was 3 years.

### Distribution of cases by gender

Our series consisted of 62 boys (60.78%) and 40 girls (39.22%) with a sex ratio M/F of 1.55.

**Figure 2.**
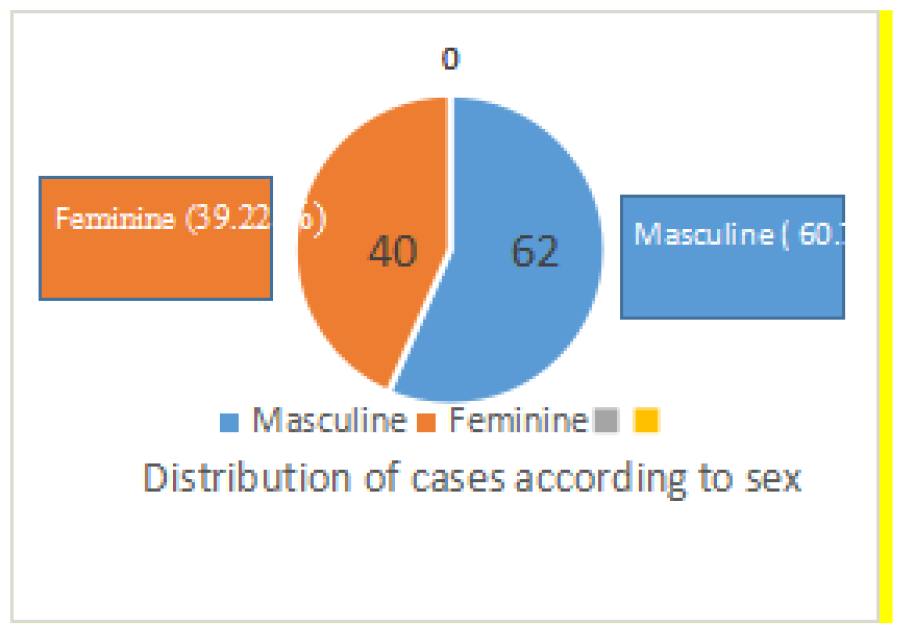
Distribution of cases according to sex.

### Distribution of cases by age group and sex

The male sex predominated in all age groups except in the over 14s where there were as many boys as girls.

**Figure 3.**
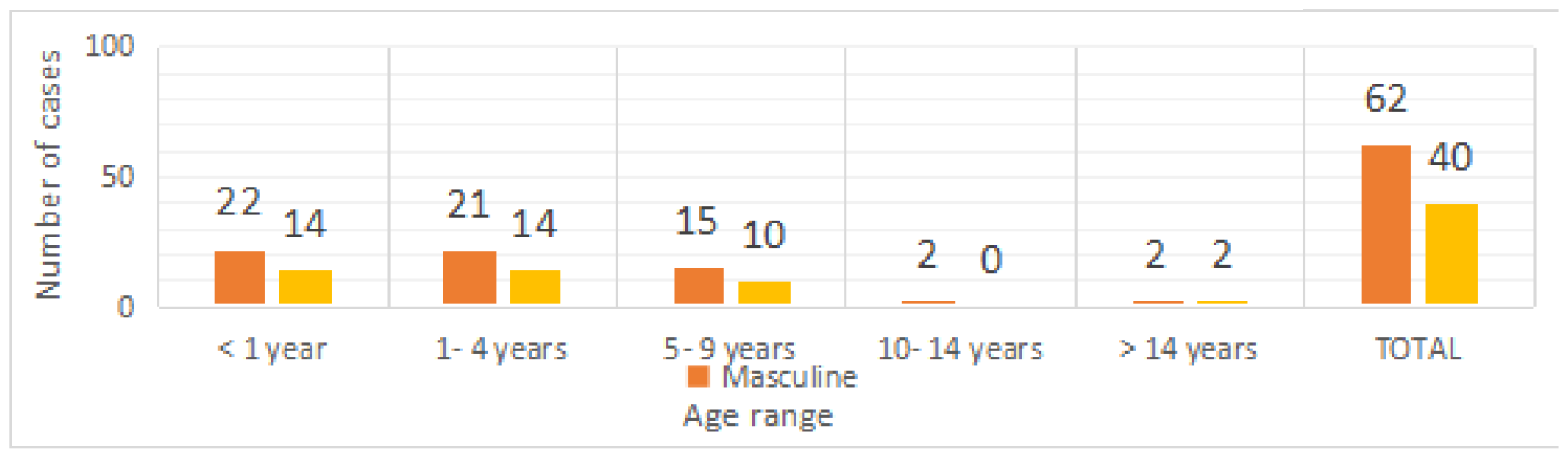
Distribution of cases by age group and sex.

### Distribution of cases by city of origin

The majority of patients came from the West department and no patient from the Northeast was recorded. The cities of origin of the other patients are presented in table 2

**Table 1.**
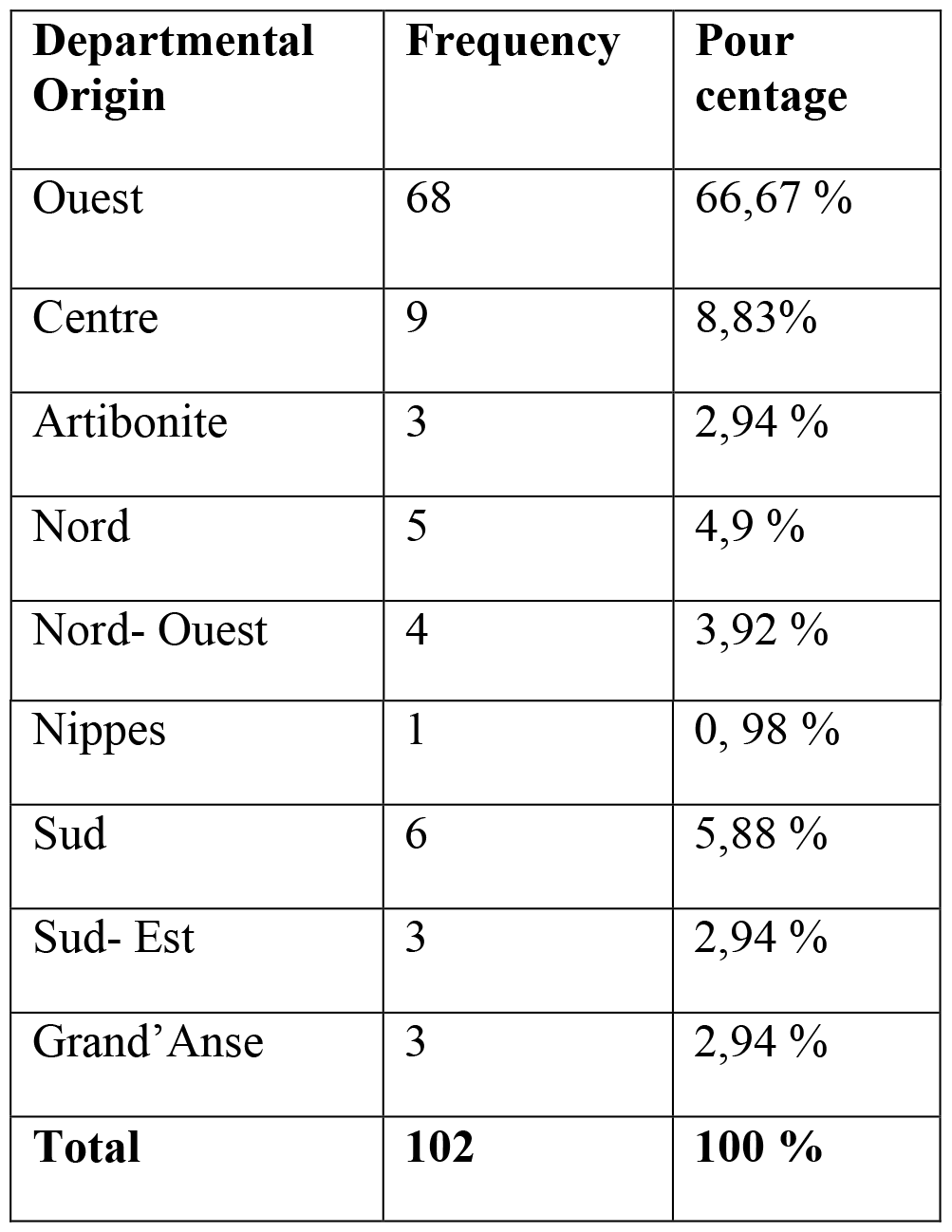
Distribution of cases according to city of origin.

| Departmental Origin | Frequency | Pourcentage |
| --- | --- | --- |
| Ouest | 68 | 66,67 % |
| Centre | 9 | 8,83% |
| Artibonite | 3 | 2,94 % |
| Nord | 5 | 4,9 % |
| Nord- Ouest | 4 | 3,92 % |
| Nippes | 1 | 0,98 % |
| Sud | 6 | 5,88 % |
| Sud- Est | 3 | 2,94 % |
| Grand'Anse | 3 | 2,94 % |
| **Total** | **102** | **100 %** |

### Description of clinical aspects

According to the body mass index measurement, growth and weight retardation was noted in 45.10% of patients. 4% of operated patients and 9% of patients with mild and moderate stenosis had a delay in growth.

**Figure 4.**
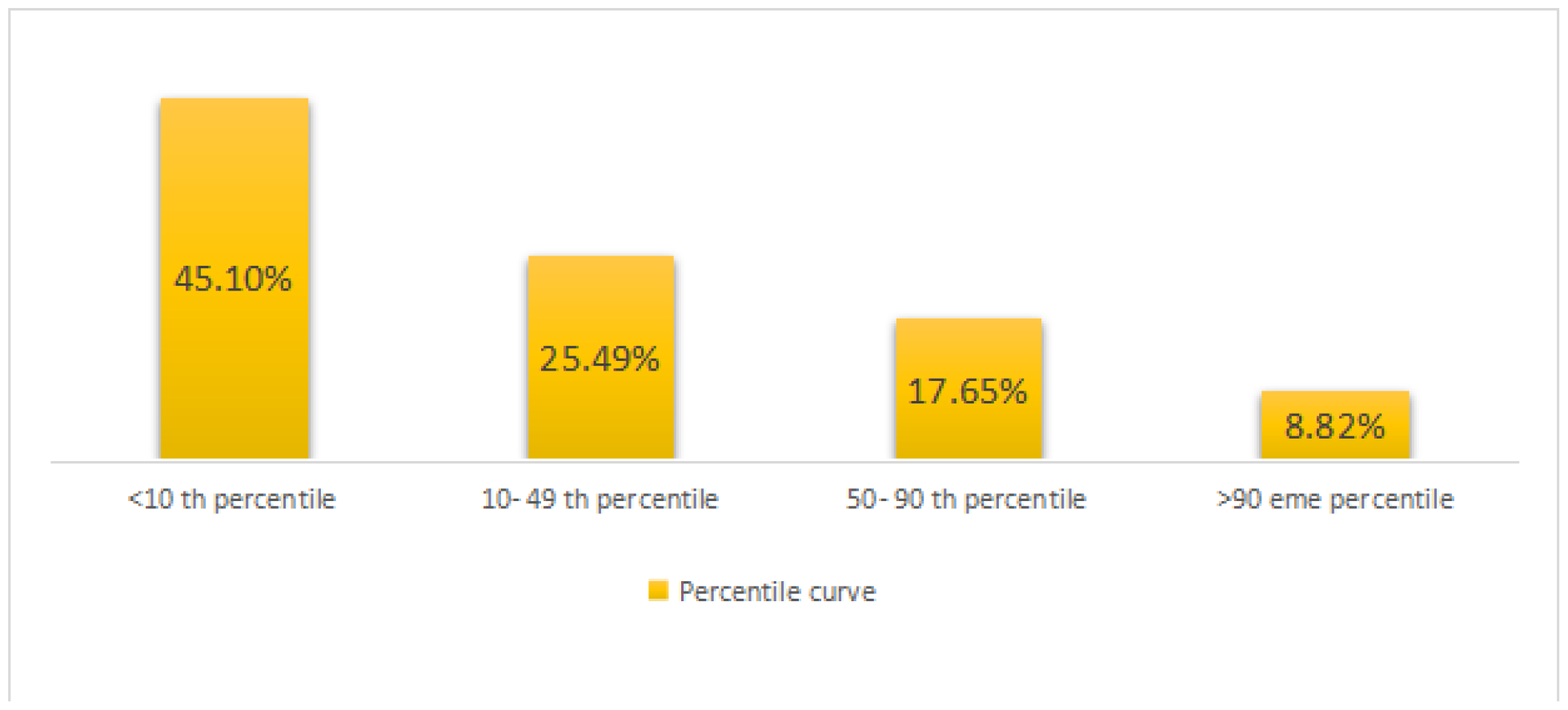
Distribution of cases according to body mass index on the percentile curve.

**Figure 5.**
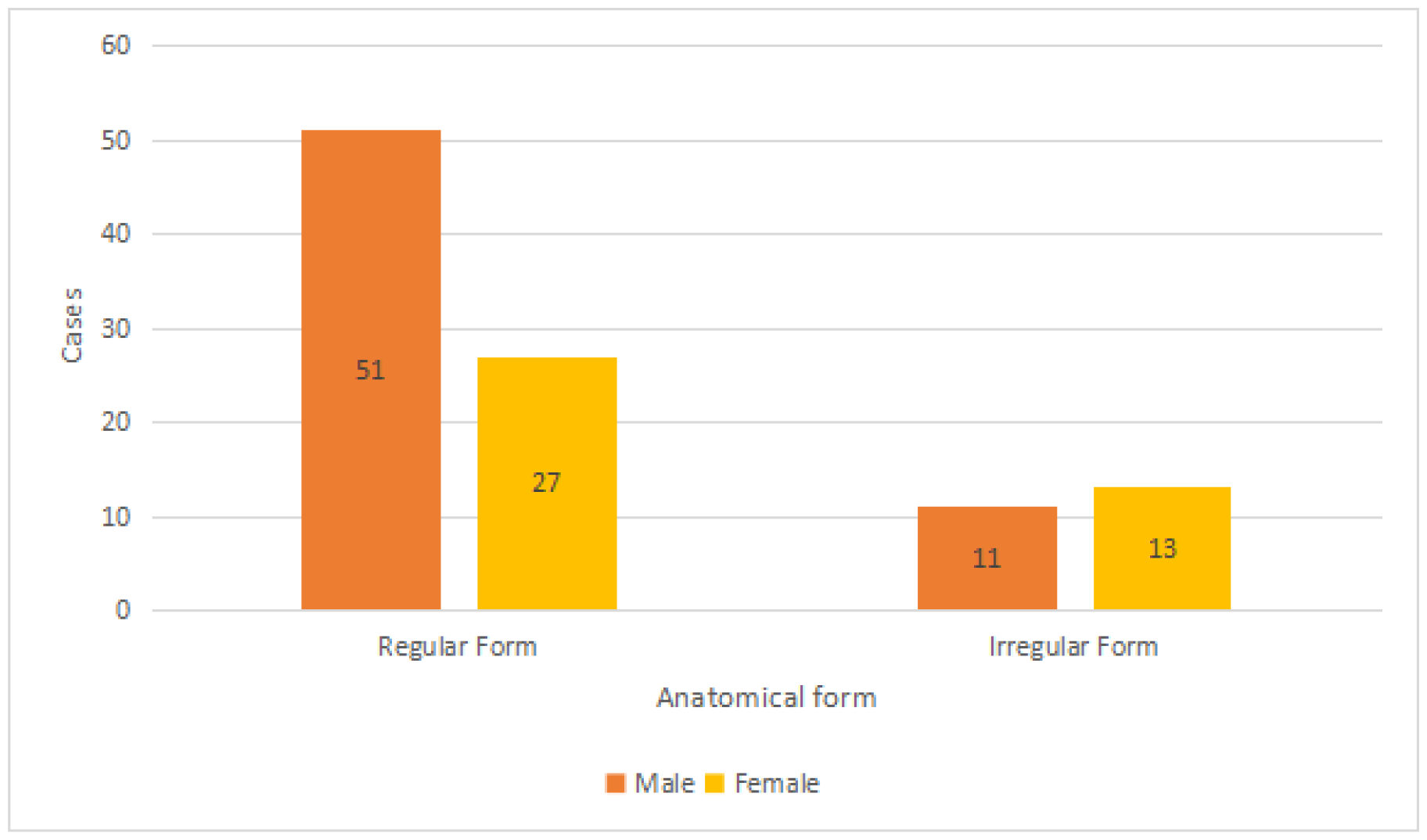
Distribution of cases according to sex and anatomical form.

The majority of patients had a severe degree of stenosis, i.e. 83.33% of cases.

The clinical signs were mostly marked in severe stenosis.

**Figure 6.**
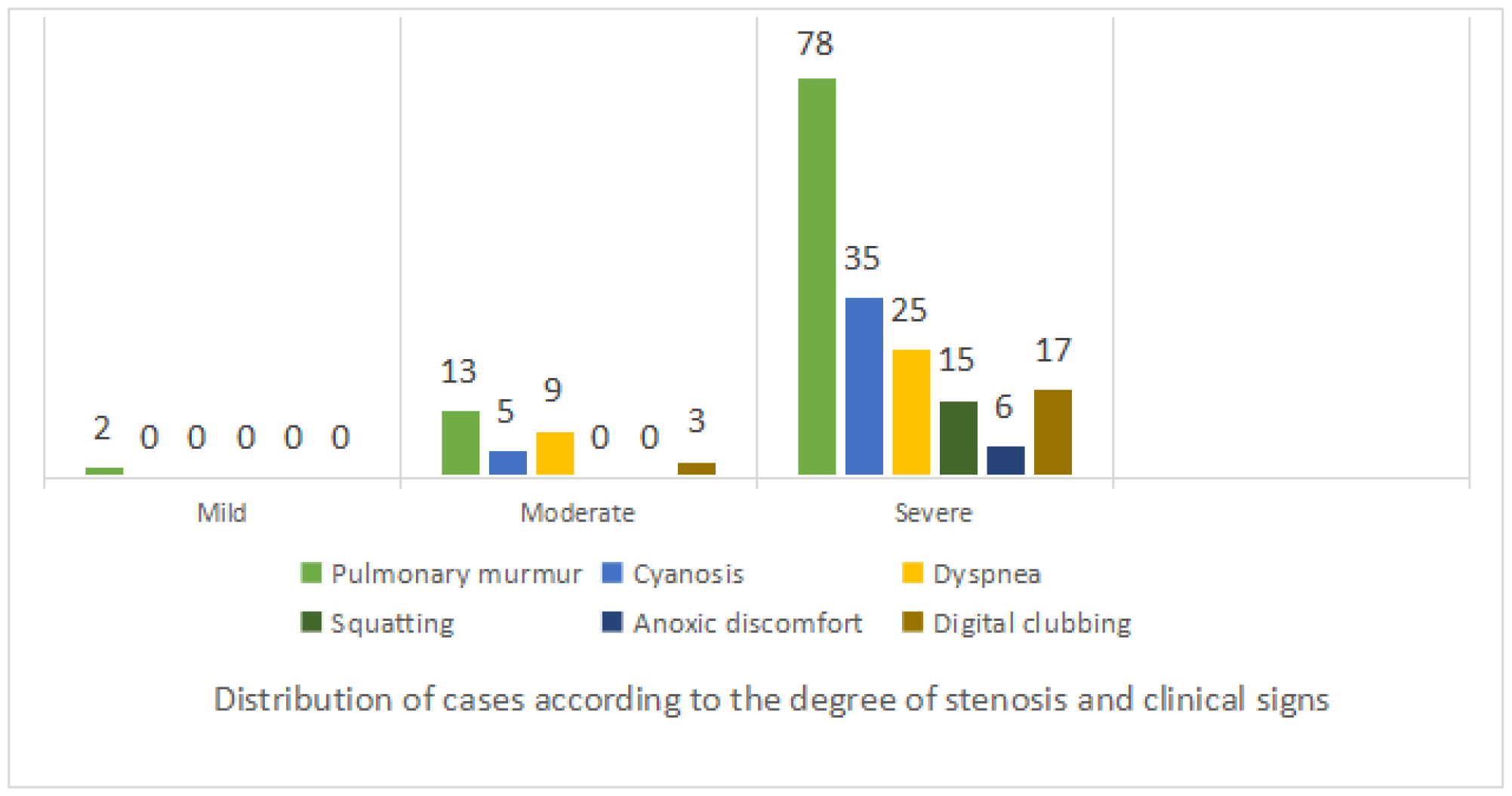
Distribution of cases according to the degree of stenosis and clinical signs.

**Figure 7.**
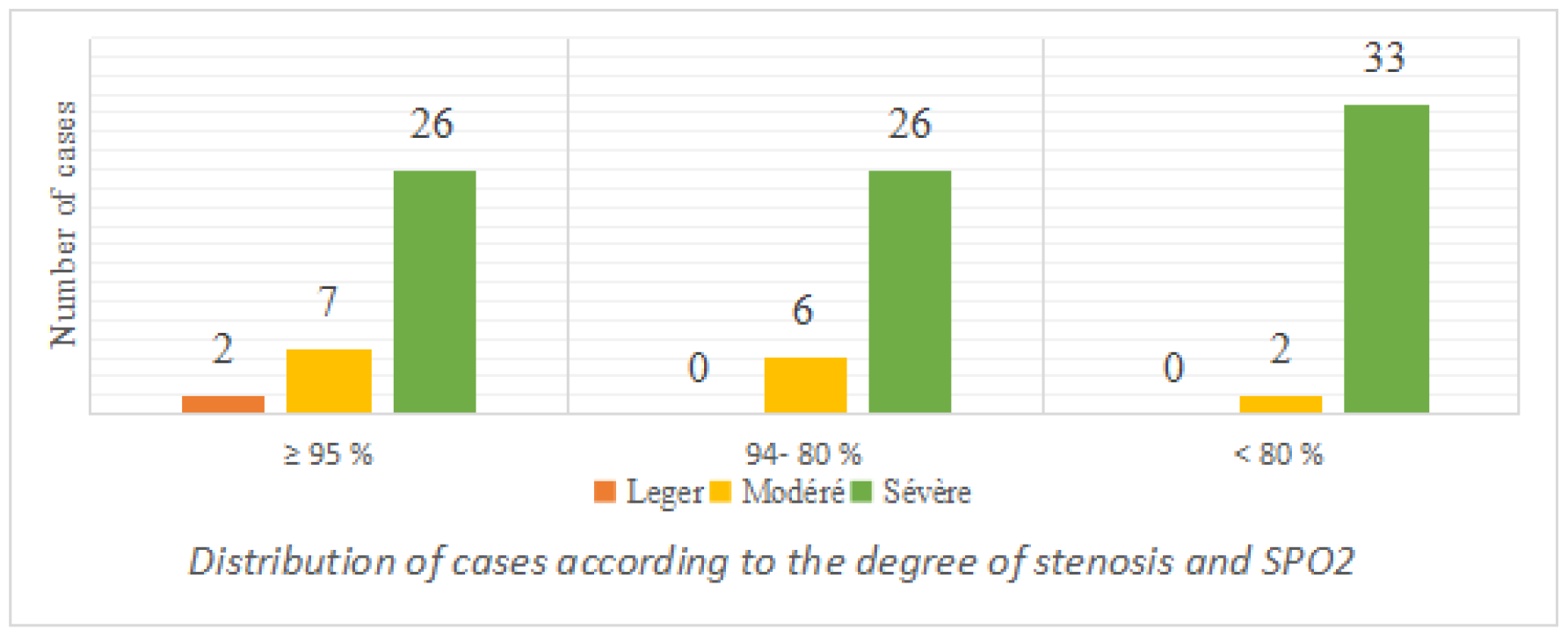
Distribution of cases according to the degree of stenosis and pulse oxygen saturation.

### Descriptions of evolving aspects

The mortality rate in the series was 14.94%.

The majority of deaths had a severe degree of stenosis.

**Figure 8.**
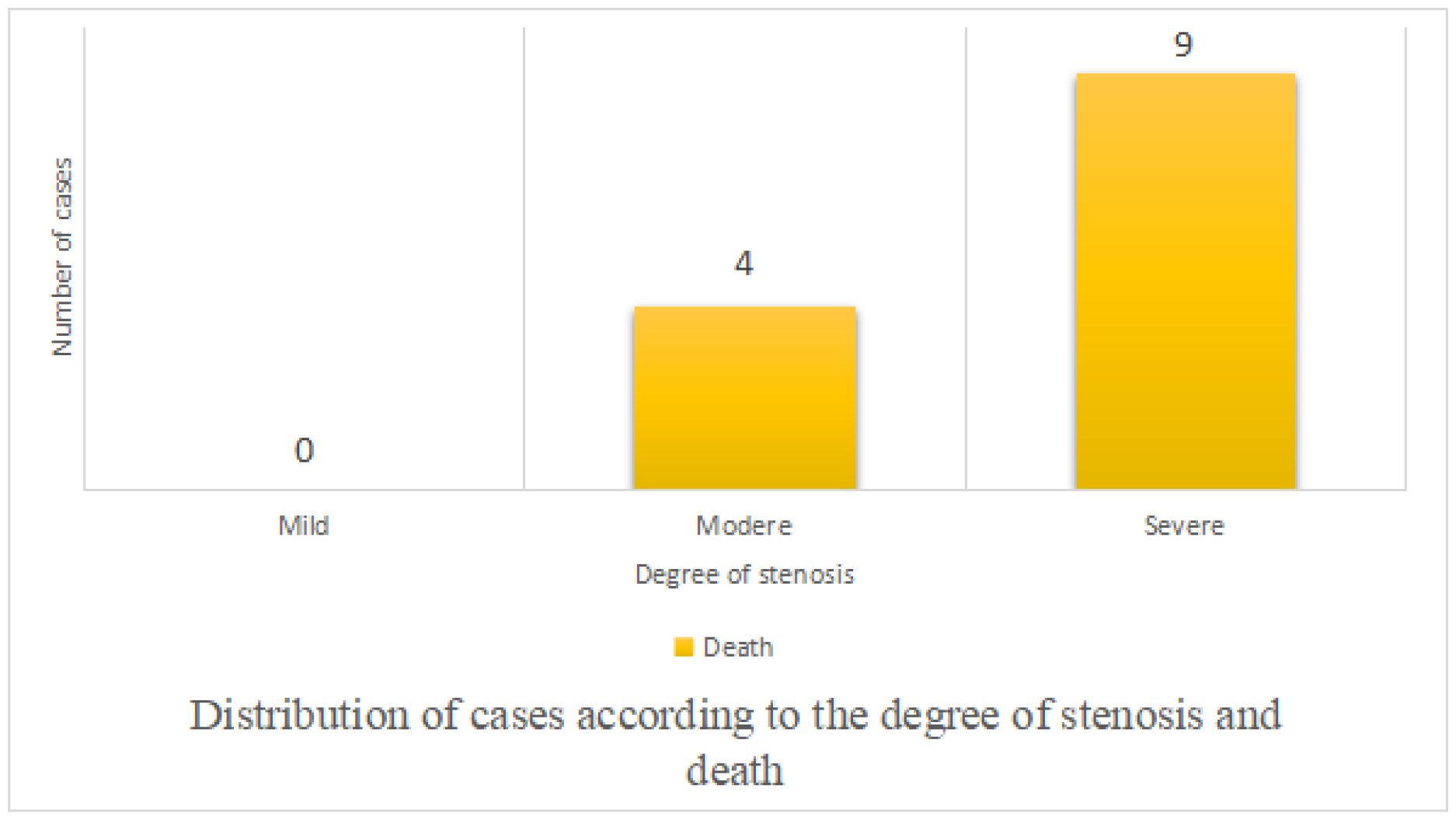
Distribution of cases according to the degree of stenosis and death.

### Description of therapeutic aspects

The majority of patients were placed on propranolol. The 6 patients who received martial treatment were under 5 years old. Iron treatment was not systematic given due to the unavailability of iron assessment at the hospital. Those with mild stenosis and those who had previously undergone surgery were without medication.

Frequency and percentage of patients treated with propranolol is 76 patients (74.51%), iron is 6 patients (5.88%) and with aspirin is 1 patient (0.98%). Percentage of patients treated surgically is 20.59% of patients (n= 21) in the series. These interventions were carried out in the United States.

The average age of patients who received surgical treatment was 5.81 years.

## 4. Discussion

Among patients aged 0 to 18 years old, there were 102 cases, representing a prevalence of 5.1% for our period. The stable drop in patients encountered during 2020 and 2021 was due first to the restrictions placed on the hospital due to the occurrence of the Coronavirus epidemic in March 2020, followed by the socio-political instability which only gets worse day by day. This has significantly reduced the influx of patients. The prevalence found in the study carried out by The Haiti cardiac alliance experience (5.59%) corroborates our results (3). Our prevalence is similar to other studies except for the study done in Burkina Faso (monocenter) where the prevalence is high (4).

All ages combined, patients aged less than five years were part of the group where the most cases of TF were diagnosed, i.e. 69.61% of cases. Our results contrast more with those of Kinda et al (1-30 months) (4). The average age of our series was 3.7 years. Our data is similar to that found in the study by M. Abdelmalik (2 years and 6 months) (5) and that found in the study by Akoudad et al (2 years and 2 months) (6). These last 2 studies carried out in Morocco as well as Haiti and Burkina Faso are part of the group of low-middle income countries, which explains the advanced age of discovery. The average age of discovery in our cohort is very high, which gives time for complications to set in. These complications are the very reasons for consultation for most. Which allows us to say that the children reached an advanced stage of the pathology at their first doctor appointment.

Concerning the gender, the majority of our patients were boys with a sex ratio of 1.55. This predominance has been noted by other authors except for the series by G. Kinda et al (4).

Concerning the degree of stenosis, seen on echocardiography, severe stenosis represented 83.33% of cases. Which is clearly higher than the series of M. Abdelmalik (65%) (5) but lower than that of A. Khayat (93.54%) (7). Therapeutic aspect: Propranolol was prescribed at 74.51% which corresponds to the series of Alexiou et al (75%) (8) and lower than the series of M. Abdelmalik (87%) (5), A. Khayat (93, 55%) (7). The iron treatment and aspirin prescribed to patients is inferior to other series. Surgical treatment was carried out in 20.59% of patients with an average age of 5.81 years while in the cohort of T. Karl et al the average age at surgical repair was 15.3 months (9). The average age is close to M. Abdelmalik’s series (5) but is higher than other studies.

The high mortality rate in our cohort can be explained by the absence of cardiac surgery in the country. Most of these surgeries were performed outside the country and are made possible with funds from international organizations. The high cost of these interventions and travel are also not covered by the Haitian state. Boundaries: As the study was descriptive and retrospective, the only sources of information were patient records and the institution’s database. Some files could not be used because they were not found in the hospital archives. The death rate was underestimated because most deaths occurred outside the hospital setting. Recommendations with a view to improving the diagnosis and management of Tetralogy of Fallot in Haiti, our recommendations are as follows: At the Ministry of Public Health and Population, - Implement an antenatal diagnosis system. - Establish social security coverage for any child suffering from congenital malformations. - Enable accessibility to care and screening for heart disease in both urban and rural areas. - Promote the training of qualified personnel in pediatric cardiology and cardiac surgery across the country. Strengthen the education of the population through awareness campaigns. To health professionals, - Encourage early detection of congenital heart disease. - Educate patients diagnosed with Tetralogy of Fallot. Refer patients who require specialized care or any patients who need it in a timely manner

## 5. Conclusion

Tetralogy of Fallot is one of the causes of infant mortality in developing countries. Haiti is no exception to this reality where this pathology is underdiagnosed. This research work is a retrospective and descriptive study of 102 patients with Tetralogy of Fallot seen at the cardiology clinic of Saint-Damien hospital from January 2019 to December 2021. The objectives consisted of the epidemiological and clinical evaluation of Tetralogy of Fallot in a pediatric population aged 0 to 18 years. Among the heart diseases found in the cardiology clinic, Tetralogy of Fallot was not negligible as a diagnosis in our cohort. The average age of discovery was late with a male predominance. Serving the entire national territory, these children mainly came from the western department. The clinical signs that predominated during the first consultation were the following: a systolic heart murmur at the pulmonary focus, followed by cyanosis and dyspnea. The regular form was the most encountered associated with a severe degree of stenosis at the time of diagnosis. Propranolol was the medical treatment of choice. Surgical treatment was indicated in all patients but only a quarter of patients were able to undergo surgery at an advanced age. The survival age was low with a high mortality rate. This work was able to confirm that TF is a reality and was able to demonstrate that the children come at the clinic at an advanced stage and therefore treatment was delayed with a high mortality rate. The findings were similar to studies carried out in developing countries. Hoping that this study will be considered a pillar in the establishment of effective epidemiological surveillance of TF within the Haitian population and will prompt other in-depth studies on the therapy and evolution of TF.

## Data Availability

All data produced in the present study are available upon reasonable request to the authors

## 7. Consideration of ethics

On November 26, 2021, we obtained authorization from our faculty, Faculty of Medicine and Health Sciences of the University of Notre Dame D’Haïti (FMSS-UNDH) to carry out the survey. We obtained formal authorization from the research department of Saint-Damien hospital on February 7, 2022 and the investigation began on February 18, 2022.

## 8. Authorship Contributions

✓ **Naïka Paulemie Désir** participated in conception and design of the study, data acquisition, data analysis, manuscript preparation and editing.
✓ **Adonaï Aly Isaac Julien** participated in data analysis, manuscript preparation, critical
✓ **Richcard Alexandre** participated in manuscript preparation, critical review and editing of the finalizing manuscript.
✓ **Joanne Fevry** participated in manuscript preparation, critical review and editing of the finalizing manuscript.
✓ **Taïna Brice** participated in manuscript preparation, critical review and editing of the finalizing manuscript.
✓ **Alexandra Noisette** participated in manuscript preparation, critical review and editing of the finalizing manuscript .

## 9. Acknowledgements

We would like to thank the staff of the Cardiology Clinic department at Saint Damien Hospital who allowed us to carry out this work.

